# SARS-CoV-2 infection in pregnancy in Denmark – characteristics and outcomes after confirmed infection in pregnancy: a nationwide, prospective, population-based cohort study

**DOI:** 10.1101/2021.06.08.21258480

**Authors:** Anna JM Aabakke, Lone Krebs, Tanja G Petersen, Frank S Kjeldsen, Giulia Corn, Karen Wøjdemann, Mette H Ibsen, Fjola Jonsdottir, Elisabeth Rønneberg, Charlotte S Andersen, Iben Sundtoft, Tine Clausen, Julie Milbak, Lars Burmester, Birgitte Lindved, Annette Thorsen-Meyer, Mohammed R Khalil, Birgitte Henriksen, Lisbeth Jønsson, Lise LT Andersen, Kamilla K Karlsen, Monica L Pedersen, Åse Klemmensen, Marianne Vestgaard, Dorthe Thisted, Manrinder K Tatla, Line S Andersen, Anne-Line Brülle, Arense Gulbech, Charlotte B Andersson, Richard Farlie, Lea Hansen, Lone Hvidman, Anne N Sørensen, Sidsel L Rathcke, Katrine H Rubin, Lone K Petersen, Jan S Jørgensen, Lonny Stokholm, Mette Bliddal

## Abstract

**Introduction:** Assessing the risk factors for and consequences of infection with SARS-CoV-2 during pregnancy is essential to guide clinical guidelines and care. Previous studies on the influence of SARS-CoV-2 infection in pregnancy have been among hospitalised patients, which may have exaggerated risk estimates of severe outcomes because all cases of SARS-CoV-2 infection in the pregnant population were not included. The objectives of this study were to identify risk factors for and outcomes after SARS-CoV-2 infection in pregnancy independent of severity of infection in a universally tested population, and to identify risk factors for and outcomes after severe infection requiring hospital admission.

**Material and Methods:** This was a prospective population-based cohort study in Denmark using data from the Danish National Patient Register and Danish Microbiology Database and prospectively registered data from medical records. We included all pregnancies between March 1 and October 31, 2020 and compared women with a positive SARS-CoV-2 test during pregnancy to non-infected pregnant women. Cases of SARS-CoV-2 infection in pregnancy were both identified prospectively and through register linkage to secure that all cases were identified and that cases were pregnant during infection. Main outcome measures were pregnancy, delivery, maternal, and neonatal outcomes. Severe infection was defined as hospital admission due to COVID-19.

**Results:** Among 82 682 pregnancies, 418 women had SARS-CoV-2 infection during pregnancy, corresponding to an incidence of 5.1 per 1000 pregnancies, 23 (5.5%) of which required hospital admission due to COVID-19. Risk factors for infection were asthma (OR 2.19 [1.41–3.41]) and being foreign born (OR 2.12 [1.70–2.64]). Risk factors for hospital admission due to COVID-19 included obesity (OR 2.74 [1.00–7.51]), smoking (OR 4.69 [1.58–13.90]), infection after gestational age (GA) 22 weeks (GA 22–27 weeks: OR 3.77 [1.16–12.29]; GA 28–36 weeks: OR 4.76 [1.60–14.12]) and having asthma (OR 4.53 [1.39–14.79]). We found no difference in any obstetric or neonatal outcomes.

**Conclusions:** Only 1 in 20 women with SARS-CoV-2 infection during pregnancy require admission to hospital due to COVID-19. And severe outcomes of SARS-CoV-2 infection in pregnancy are rare.

**Key Message:** Population based cohort study about SARS-CoV-2 infection during pregnancy. Asthma and foreign ethnicity were identified as risk factors for infection while obstetric outcomes did not change. Obesity, smoking, infection after GA 22, and asthma increased the risk of hospital admission.

## Main body of Text

### Introduction

The World Health Organisation declared a global pandemic of coronavirus disease (COVID-19) in March 2020.^1^ COVID-19 is caused by infection with severe acute respiratory syndrome coronavirus 2 (SARS-CoV-2).

Assessing the risk factors for and consequences of infection with SARS-CoV-2 during pregnancy is essential to guide clinical guidelines and care.

Evidence indicates that pregnant women are not at increased risk of infection with SARS-CoV-2 compared to non-pregnant women.^2–4^ Risk factors associated with infection include obesity, age, ethnicity, and pre-existing morbidities, and SARS-CoV-2 infection in pregnancy has been associated with an increased risk of admission to an intensive care unit (ICU), preterm delivery, and admission of the neonate to a neonatal intensive care unit (NICU).^5–9^

Most previous population-based studies on SARS-CoV-2 infection in pregnancy have been among hospitalised patients, which may have exaggerated risk estimates of severe outcomes because all cases of SARS-CoV-2 infection in the pregnant population were not included.^6,10^ Only one study from the Netherlands included all cases independent of severity.^7^ However, in that study cases were identified prospectively in obstetric clinics, with the risk of selection bias. Additionally, the outcomes are in most studies compared to outcomes of historic cohorts of non-infected women, with the risk that the consequences of the pandemic will be attributed solely to SARS-CoV-2.^6,7,11^ Lastly, testing strategies influence the estimated prevalence of SARS-CoV-2 infection, and potentially also the estimated severity of outcomes in that also the mild and asymptomatic cases detected.^5^

The testing strategy in Denmark rapidly evolved during the pandemic from testing the most ill in March 2020 to testing cases with mild symptoms from April and testing close contacts from May. Universal testing of all pregnant women admitted to hospital including those for delivery was implemented nationwide in early May 2020.

The objectives of this study were to identify risk factors for SARS-CoV-2 infection in pregnancy in a universally tested population and risk factors for severe infection requiring hospital admission, and to investigate the consequences of infection and severe infection on pregnancy, delivery, and neonatal outcomes when comparing to all non-infected pregnancies during the same time period.

## Material and methods

This was a nationwide prospective population-based study investigating the association between SARS-CoV-2 infection in pregnancy and clinical characteristics and maternal, delivery, and neonatal outcomes. The study used prospectively registered data from medical records registered in the Danish COVID-19 in pregnancy database (DCOD) and register data obtained from the following national registers: the Danish National Patient Register (DNPR),^12^ Danish Microbiology Database (MiBa),^13^ and the Civil Registration System.^14^

The overall study population was identified in DNPR and comprised all women registered with a pregnancy or birth-related ICD10 diagnosis or procedure between March 1 and October 31, 2020 as specified in Table S1. SARS-CoV-2-positive cases within the study population were identified by linkage to MiBa. In Denmark, all positive polymerase chain reaction (PCR) tests are registered in MiBa, while antigen and antibody tests were not during the study period. Pregnant women identified in the registers were followed up until December 11, 2020.

In the DCOD, women with a positive SARS-CoV-2 test during pregnancy between March 1 and October 31, 2020 were registered prospectively. Eligible SARS-CoV-2 tests included PCR tests (of pharyngeal swab or tracheal secretion), antigen tests (of nasal swab), or detection of antibodies (in a blood test) combined with a history of COVID-19 symptoms during pregnancy. All obstetric units in Denmark participated, and clinicians from each unit reported cases based on data in the medical records. DCOD was based in EasyTrial (easytrial.net, Denmark). Cases were validated every second month by register linkage, with data obtained from the DNPR, Danish National Service Register, and MiBa. The validation dataset included only personal identification number, date of positive SARS-CoV-2 PCR test, date of hospital contact, and name of the hospital. Non-reported cases identified by validation who were pregnant at the time of a positive SARS-CoV-2 PCR test were entered into the DCOD retrospectively. Women included in the DCOD were followed up until data extraction on February 8, 2021.

The register data on the overall study population were pseudoanonymised, making it impossible to validate the SARS-CoV-2-positive cases from the registers against the DCOD cases on an individual level. We therefore present data for SARS-CoV-2-positive women from the DCOD alongside cases identified in the registers to illustrate degree of comparability. SARS-CoV-2 cases in the DCOD and those registered in the national registries were each compared to the non-infected population. The validity of maternal and neonatal characteristics, surgical procedures, and main diagnoses in the Danish registers is considered high,^15^ and we therefore evaluated that a comparison between the DCOD data and the registry data was relevant.

The estimated date of delivery (EDD) was registered in the DCOD and based on the first trimester ultrasound examination offered to all pregnant women between gestational age (GA) 11 and 14 completed weeks (w). In case of first trimester abortion, the EDD was calculated based on the GA at abortion. In the DCOD, date of the first day of the last menstrual period (LMP) was defined as EDD minus 280 days. In the register data, the LMP was calculated based on the registered GA at delivery or abortion. If still pregnant, we calculated the LMP based on GA at ultrasound examination, and if missing, the recommended GA for the registered ultrasound examinations. For abortions with a missing registration of GA, GA was imputed based on the mean GA within the categories miscarriage or induced abortion.

Characteristics included maternal age at LMP, body mass index (BMI) based on pre-pregnancy weight as registered at first contact in pregnancy, smoking defined as any smoking during pregnancy, parity, multiple pregnancy, pre-eclampsia, gestational diabetes, and diabetes. The register dataset also included pre-existing asthma, and hypertension diagnosed within 5 years of LMP. Table S2 presents the codes used to identify characteristics and outcomes in DNPR. The DCOD also included information on migrant status defined as not born in Denmark, employment status including ongoing education, asthma defined as asthma requiring steroid inhalation, pre-existing medical problems and previous pregnancy-related complications as specified in Table S3.

Maternal outcomes included admission to an ICU, pneumonia (in the DCOD defined as confirmed by radiological examination), thromboembolic events, maternal mortality (death during pregnancy or within 7 days postpartum) and pregnancy outcomes including delivery, termination of pregnancy before GA 23 w, or miscarriage before GA 22 w.

Delivery outcomes comprised mode of delivery, induction of labour, and preterm birth before GA 37 w.

Neonatal outcomes included admission to a NICU directly from the delivery or maternity ward, stillbirth from GA 22 w, Apgar score at 5 minutes, umbilical artery pH, and neonatal death (within 28 days postpartum).

Data on migrant status and infant admission to a NICU were not available in our register data. These variables registered in the DCOD were therefore compared to historical data comprising migrant status of all women with a liveborn child in 2019 in Denmark as reported by Statistics Denmark^16^ and admissions to a NICU of all live born children in Denmark in 2019 as reported by the Danish Quality Database for new-borns.^17^ In the DCOD, admission to hospital with a concurrent SARS-CoV-2 infection was defined as admission and discharge on two different dates and a positive SARS-CoV-2 test within 14 days before or during admission. Severe infection was defined as admission to hospital due to COVID-19 symptoms.

### Statistical Analyses

Categorical variables are presented as count with percentage, and continuous variables as median with interquartile range (IQR). For DCOD data numbers < 3 and for register data numbers < 5 are not reported to avoid identification according to the respective ethical approvals. The categorical DCOD data were analysed by calculating the odds ratio and with chi^2^ test when compared to the non-infected population, and with logistic regression when compared to severe versus non-severe DCOD cases including multivariate analyses adjusting for age, BMI, parity, and smoking. Continuous DCOD variables were analysed using the Wilcoxon signed rank test. We did a sub-analysis of the DCOD cases testing for the influence of gestational age at time of SARS-CoV-2 infection on key outcomes comprising cesarean delivery, induction of labour, preterm birth, and NICU admission. In the analysis of register data, categorical outcomes were assessed with univariate and multiple logistic regression and continuous variables with linear regression. The multivariate analyses adjusted for a) maternal age and LMP to adjust for time of inclusion in the study, and b) predefined risk factors including maternal age, LMP, parity, BMI, and smoking. In the linear regression model, we included log-transformed outcome variables, applying bootstrapping to account for non-normal distribution of residuals. For the outcomes miscarriage, termination of pregnancy, and preterm delivery, a Cox regression analysis was performed, including SARS-CoV-2 as time-dependent exposure, to account for duration of exposure to SARS-CoV-2 during follow-up. For the first three outcomes, women were included in the survival-analysis according to the time of first registered hospital contact, and for preterm delivery, women were included at GA 100 days and registered as SARS-CoV-2 exposed at the date of positive test. Missing data were excluded from all analyses. A *P*-value ≤ 0.05 was considered statistically significant. Data were analysed using Stata/MP16 (64-bit) and IBM SPSS statistics 27 (SPSS Inc).

### Ethical Approval

The study was approved by the Danish Patient Safety Authority on the April 24, 2020 (reg. no. 31-1521-252), the regional Data Protection Agency in Region Zealand on the March 23, 2020 (reg. no. REG-022-2020), and the regional Data Protection Agency in Region of Southern Denmark on April 15, 2020 (reg. no. 20/17416). The study is reported according to STROBE guidelines.^18^

## Results

Among 82 682 pregnancies in Denmark between March 1 and October 31, 2020, 420 women had a SARS-CoV-2 positive PCR-test, and 418 SARS-CoV-2 cases were confirmed as being during pregnancy through validation against medical records. Figure 1A illustrates the flow through the register-based part of the study, and Figure 1B the flow through the DCOD study.

**Figure 1.**
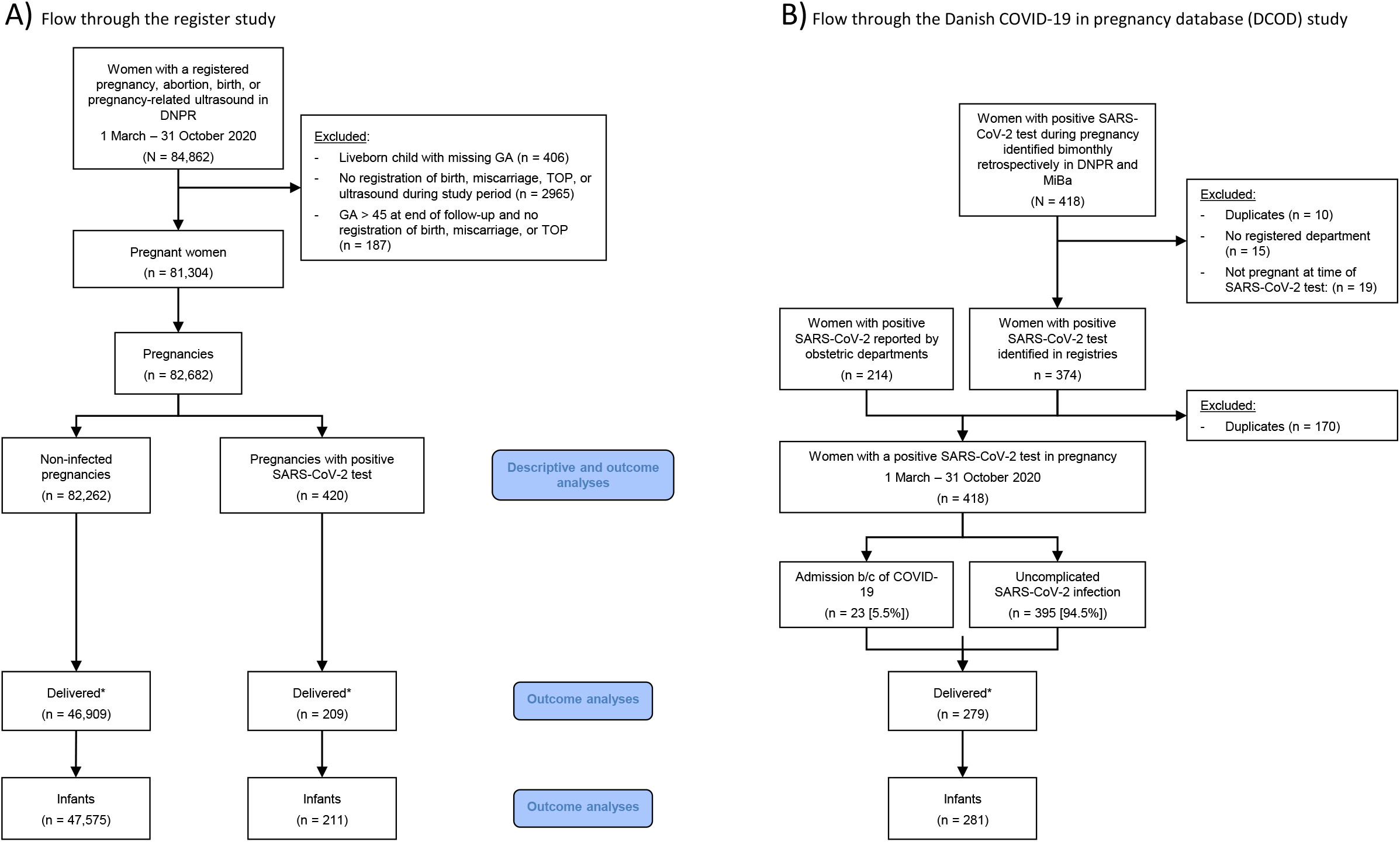
A) Flow through the register study; B) Flow through the Danish COVID-19 in pregnancy database (DCOD) study. * The number of deliveries differs in the two sub-studies due to different follow-up times. Women identified in the register study were followed up until 11 December 2020 and women identified in the DCOD study until 8 February 2021. b/c: because. DNPR: Danish National Patient Register. GA: Gestational age in completed weeks. MiBa: The Danish Microbiology Database TOP: Termination of pregnancy

The overall incidence of SARS-COV-2 infection during validated pregnancy in the inclusion period was 5.1 (95% confidence interval (CI) 4.7–5.5) per 1000 pregnancies, with the monthly incidence ranging from 0.2 per 1000 pregnancies in July to 3.8 in October (Figure S1). Of the SARS-CoV-2 cases in pregnancy, 23 (5.5%) were admitted to hospital because of severe COVID-19 symptoms.

Basic characteristics of women and their pregnancies according to SARS-CoV-2 status and severity of infection are presented in Table 1 and Table S4. Pregnant women infected with SARS-CoV-2 more frequently had pre-existing asthma (crude odds ratio (OR) 2.19, 95% CI 1.41–3.41) and were foreign born (OR 2.12, 95% CI 1.70–2.64) compared to the non-infected. There were no differences in the rates of pre-eclampsia or gestational diabetes. Women who required admission to hospital had a significantly higher BMI (*P* = 0.018), were more likely to smoke in pregnancy (OR 4.69, 95% CI 1.58–13.90), and have pre-existing asthma (OR 4.53, 95% CI 1.39–14.79). Compared to women becoming infected before, women becoming infected after GA 22 w had a higher risk of COVID-19-related hospital admission (GA 22–27 w: OR 3.77, 95% CI 1.16–12.29; GA 28–36 w: OR 4.76, 95% CI 1.60–14.12).

**Table 1.**
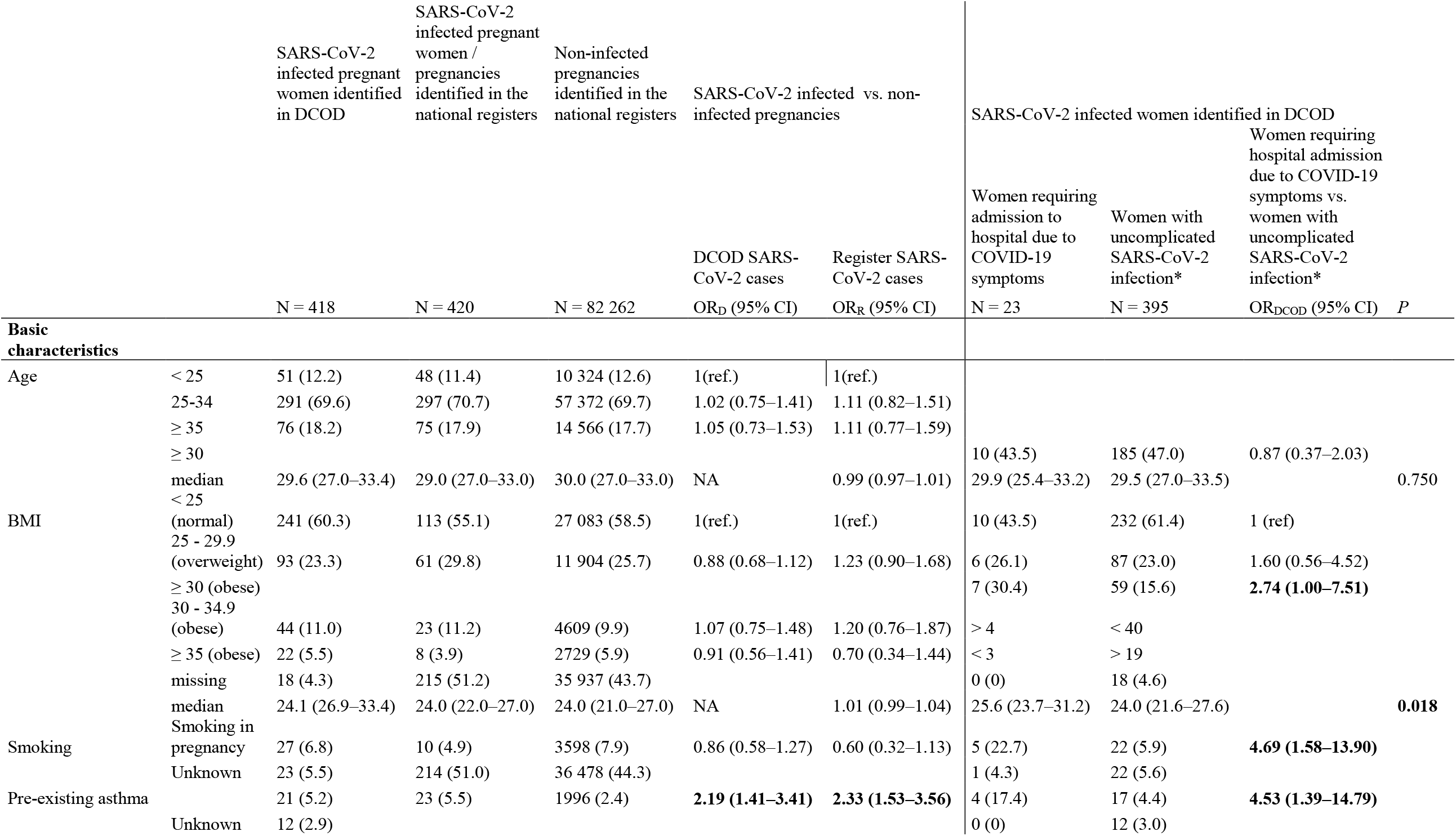
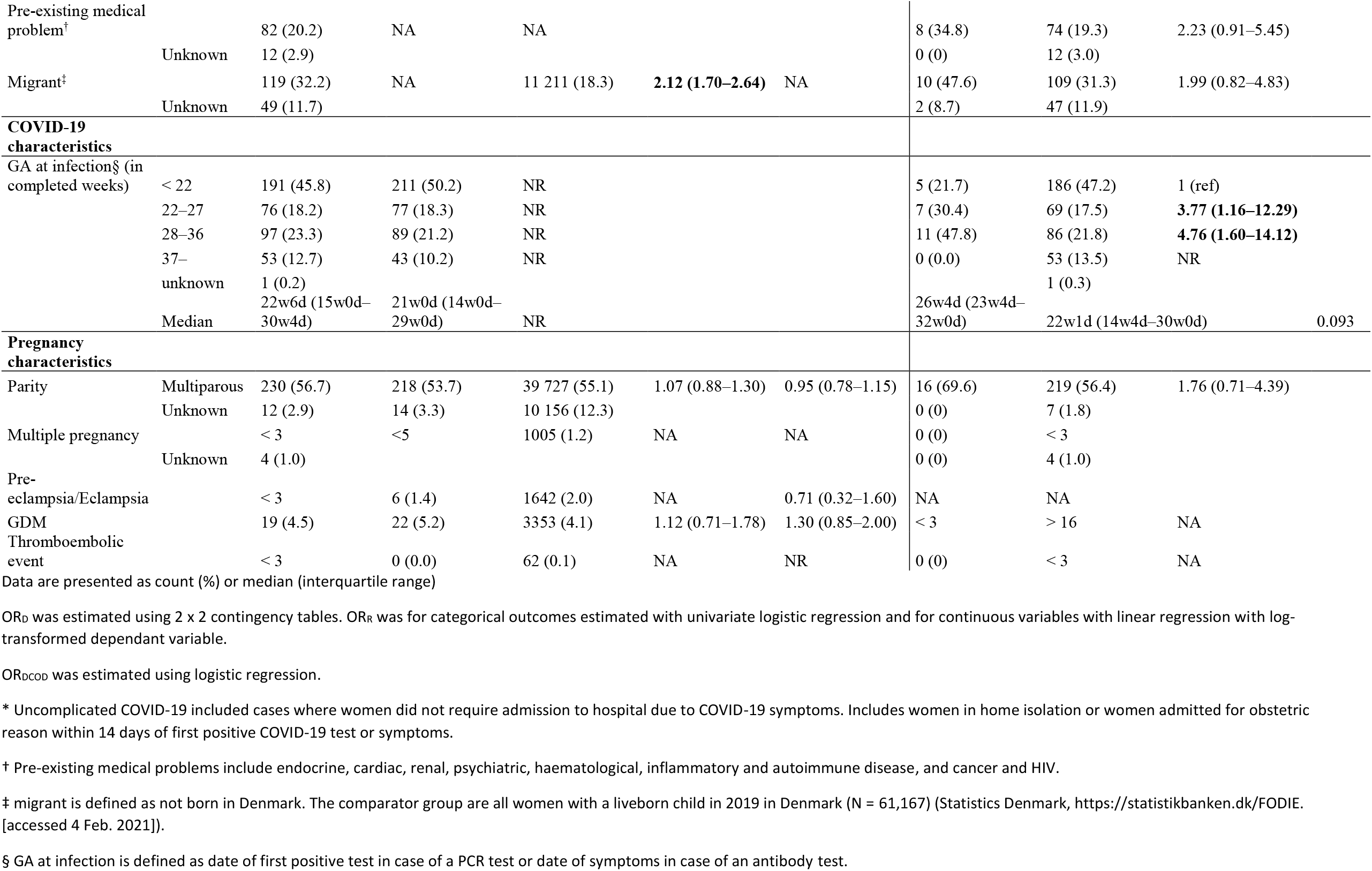
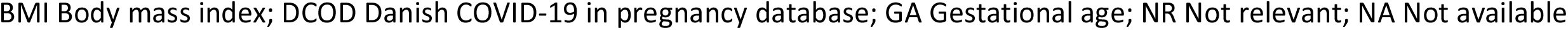
Basic characteristics of SARS-CoV-2 infected women overall and according to severity of infection and all pregnant women in Denmark between March 1 and October 31, 2020.

The outcomes of the SARS-CoV-2 infected women compared to all non-infected pregnancies are presented in Table 2. The rate of pneumonia among SARS-CoV-2 infected pregnant women was 1.5% (n = 6), corresponding to a significantly higher risk compared to non-infected pregnancies (OR 15.97, 95% CI 6.92–36.86). The rate of thromboembolic events and admission to an ICU was low, and we had no cases of maternal death during the study period. The rate of early pregnancy loss did not vary between the groups after adjusting for exposure time. However, the risk of termination of pregnancy was increased after adjustment (Hazard Ratio (HR) 2.39, 95% CI 1.29– 4.45) among the SARS-CoV-2 infected women. The median time between SARS-CoV-2 infection and delivery was 88 days (interquartile range 35–138). We found no difference between the groups regarding any of the delivery or infant outcomes. Adjustment did not change the results. We had no neonatal deaths among the children of SARS-CoV-2 infected women.

**Table 2.**
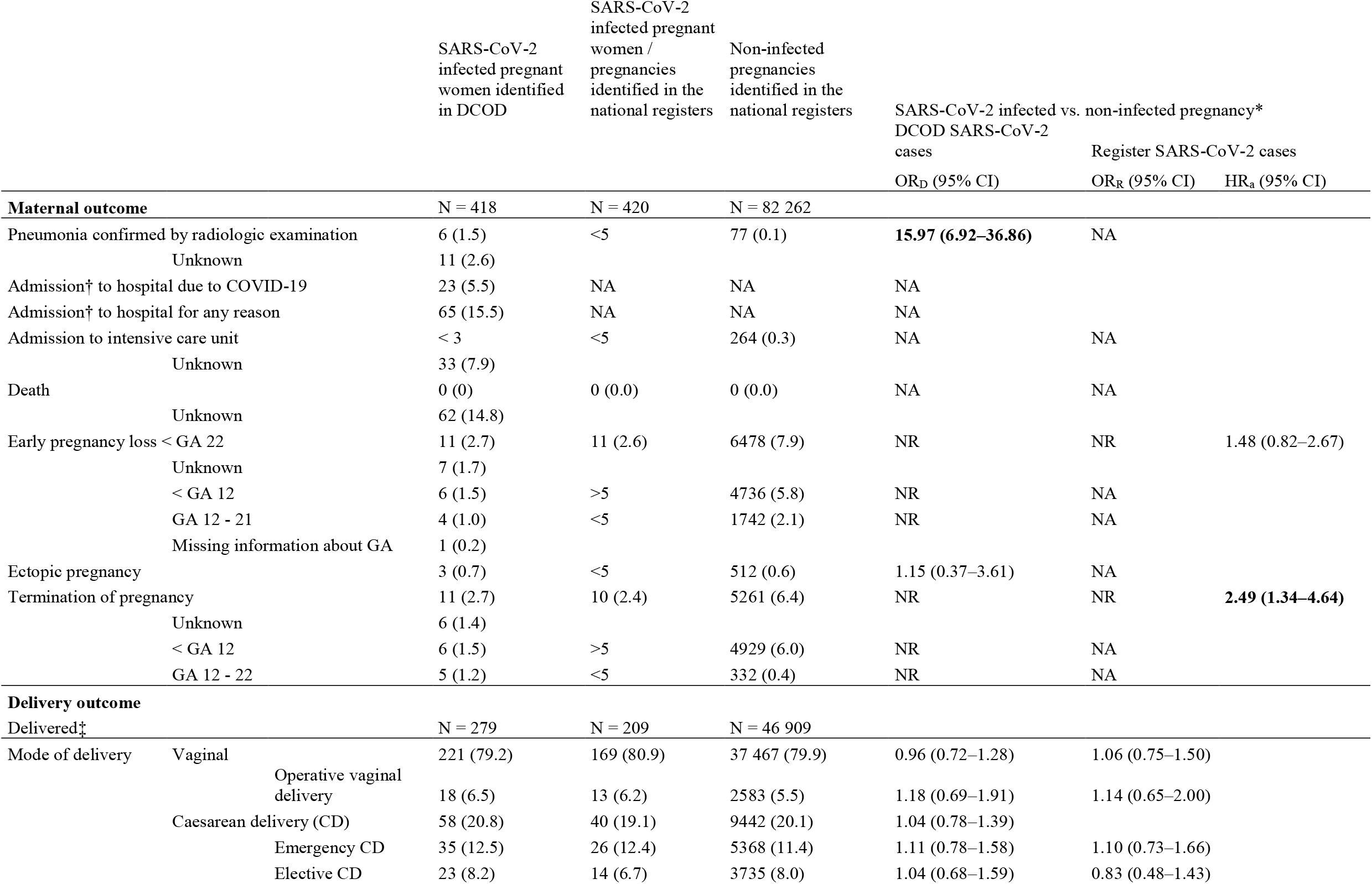
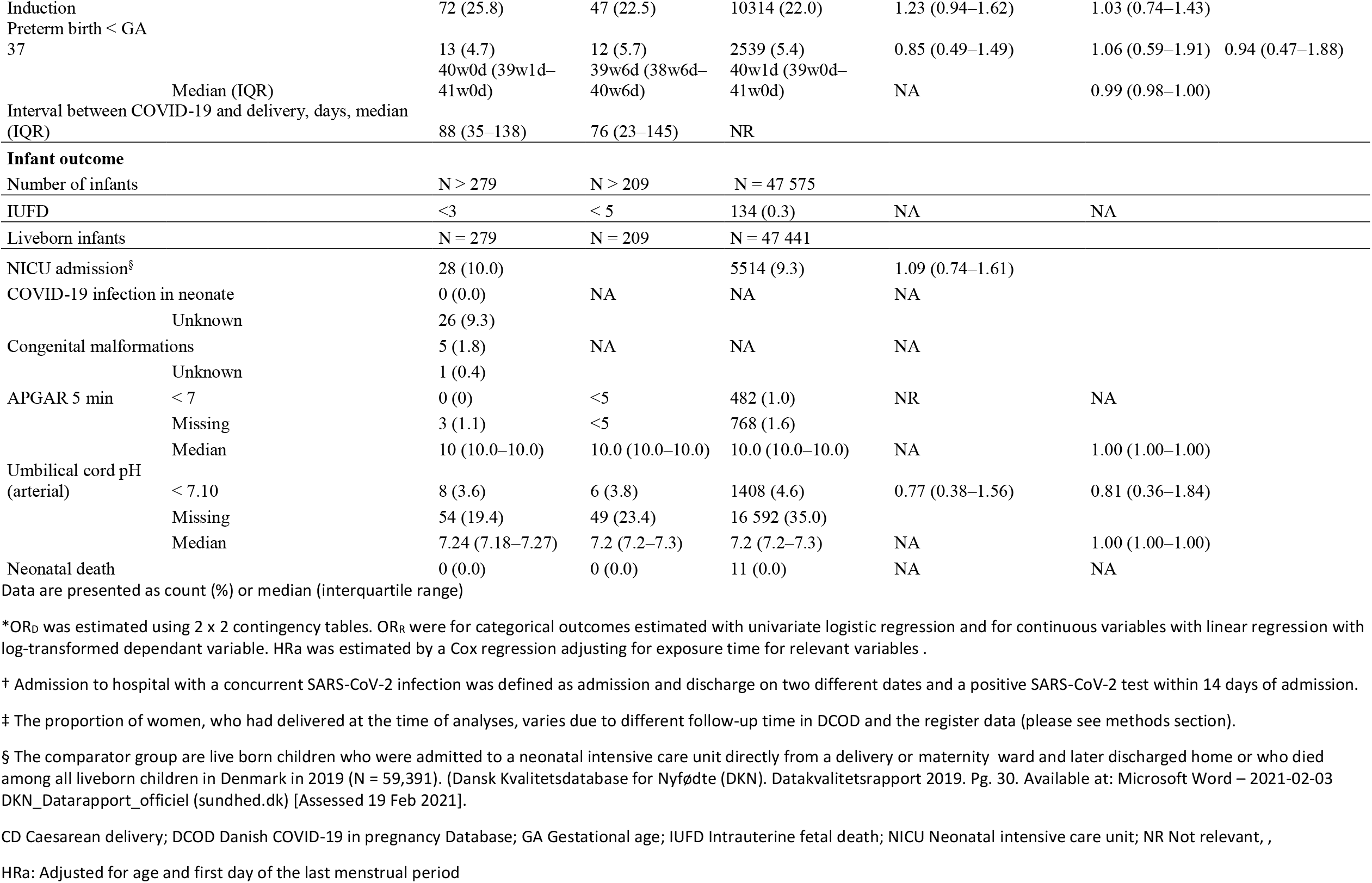
Outcomes for women with SARS-CoV-2 infection in pregnancy compared to all pregnancies in Denmark between March 1 and October 31, 2020.

We found no difference in outcomes between women with severe COVID-19 infection requiring hospital admission compared to less severe SARS-CoV-2 cases in either the crude or adjusted analyses (Table 3).

**Table 3.**
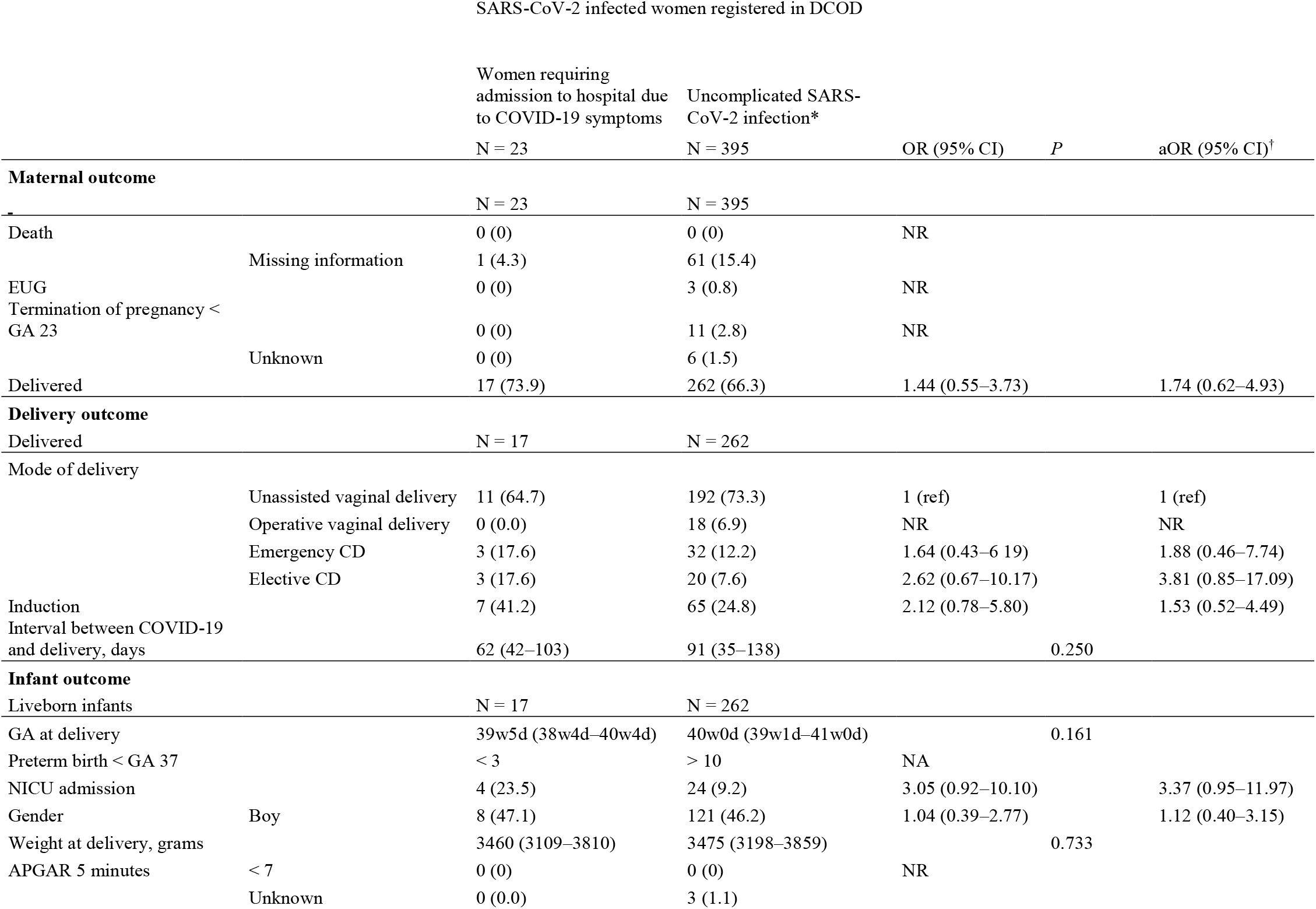
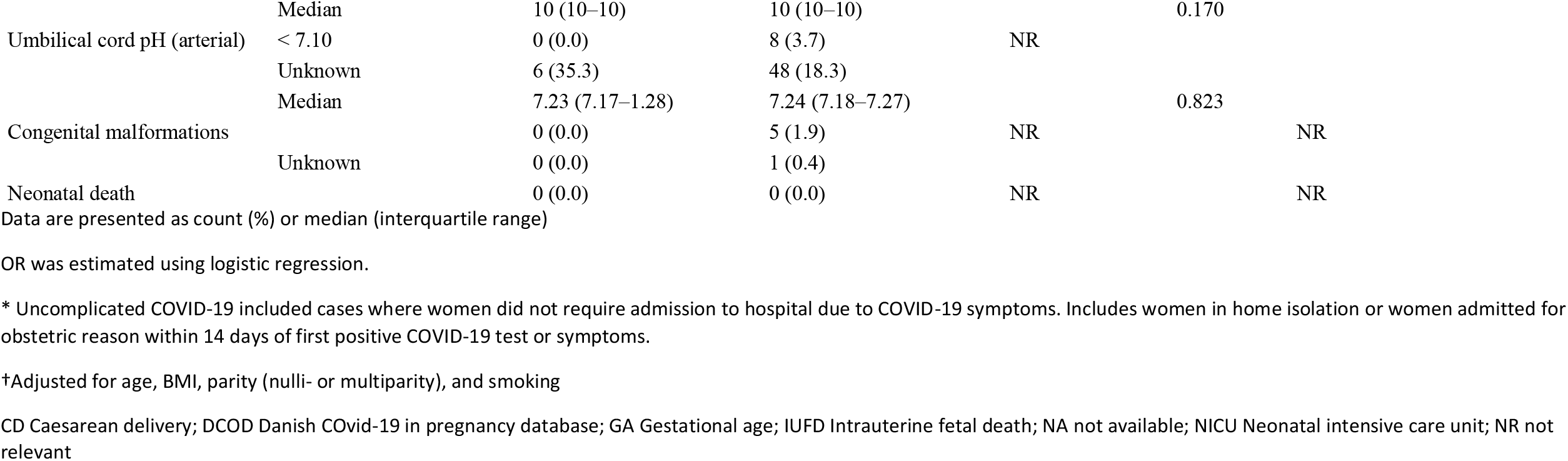
Outcomes for women with severe COVID-19 infection requiring admission to hospital compared to less severe SARS-CoV-2 cases in pregnancy in Denmark between March 1 and October 31, 2020 as registered in the Danish COVID-19 in pregnancy database (DCOD).

There was no difference in risk of cesarean delivery, induction of labour, preterm birth, or NICU admission in relation to GA at infection among the SARS-CoV-2 infected women (data not shown).

## Discussion

In this prospective population-based cohort study, we found 418 validated cases of SARS-CoV-2 among pregnant women, corresponding to an incidence of 5.1 per 1000 pregnancies. Only 23 women (5.5%) required hospital admission due to the COVID-19 infection. Risk factors for infection were asthma and being foreign born, while obesity, smoking in pregnancy, infection after GA 22 w, and having asthma increased the risk of hospital admission due to COVID-19. We found an increased risk of late termination of pregnancy, but no difference in any delivery or neonatal outcomes among infected women. Severity of infection did not affect outcomes.

Minority ethnicity has been identified as a risk factor for SARS-CoV-2 infection in both pregnant and non-pregnant women.^5–8,11^ The increased risk level found in this study is similar to that reported in a large systematic review.^5^ The aetiology of the ethnic disparity is not clear, but it is probably multifaceted and related to cultural and socioeconomic factors rather than to SARS-CoV-2.

The identified risk factors for severe infection comprising asthma, increasing maternal weight, and GA at the time of infection are similar to risk factors identified in previous studies.^5,6^ Smoking in pregnancy has not been described previously, but seems a plausible risk factor due to its effect on lung tissue. Previous population-based studies have primarily included women admitted to hospital,^6,10,11^ and our results in the more severe cases (Table 3) are therefore comparable to the results of these studies.

Women with SARS-CoV-2 infection had, expectedly, a much higher risk of pneumonia, but otherwise we found no increased risk for adverse maternal or neonatal outcomes in infected compared to non-infected pregnant women. Previous studies have found increased risks of admission to an ICU, induction of labour, preterm delivery, and admission to a NICU,^5–9,19,20^ which we could not confirm. The lack of association in our study might be related to the fact that we included all cases of SARS-CoV-2 infection in pregnancy independent of severity. The previously shown associations between SARS-CoV-2 and severe outcomes might be related to severity of disease. Albeit, we found no difference in outcomes among cases requiring hospital admission due to COVID-19 compared to less severe cases. Nevertheless, in a Nordic collaborative study, women with severe COVID-19 were at increased risk of induction of labour, cesarean delivery, and preterm delivery.^21^

We found few cases of early pregnancy complications among the SARS-CoV-2 infected women. However, after adjustment for exposure time, the risk of termination of pregnancy was higher among the SARS-CoV-2 infected women. In Denmark, all women admitted to hospital are tested for SARS-CoV-2, and we assume that the increased rate of terminations is partly related to more frequent testing and consequently a higher detection rate of SARS-CoV-2 among women opting for provoked abortion rather than the SARS-CoV-2 diagnosis itself. However, an association cannot be ruled out and future studies are needed to explore whether there is a possible association between SARS-CoV-2 infection and terminations of pregnancy.

This study has several strengths. The major strength is the combination of register-based data considered complete^12^ and prospectively collected medical record data, allowing for validation, analysis of disease severity, and multivariate analyses. Furthermore, the comparison population comprised all pregnancies from the same inclusion period as the SARS-CoV-2 cases, thus adjusting for the possible consequences of community regulations during a pandemic.^22^ Additionally, we identified the vast majority of SARS-CoV-2 cases in pregnancy because testing was widespread in Denmark during the inclusion period, and the DCOD data were validated against registry data.

The study also has limitations. First, the register dataset was pseudoanonymised, making individual-level linkage to DCOD impossible. Additionally, data on SARS-CoV-2 cases in the DCOD were based on medical records and data in the national registers were based on mandatory nationally registered data, and these data might not be directly comparable. Nevertheless, the number of SARS-CoV-19 cases was similar in the two cohorts, and data did not differ significantly between the cohorts, indicating agreement between cases and data sources. Some descriptive variables including BMI and smoking status are not reported in the registers before delivery and are not reported for early pregnancy losses. The DCOD thus provided complete data, and made national surveillance of the infection possible to support national guidelines until register data were available.^23–25^ Secondly, the lack of association in some outcomes for severe cases might be due to low numbers. Thirdly, universal testing of pregnant women was not implemented in Denmark before May 2020, and we might therefore have missed SARS-CoV-2-positive cases early in the inclusion period. Furthermore, MiBa only included information on PCR tests, thus missing pregnant women diagnosed through antigen or antibody tests, which were possibly milder cases. Inclusion of these plausibly positive but non-identified cases in the comparison population of pregnancies might have affected our estimates.

## Conclusion

In this prospective population-based cohort study with universal SARS-CoV-2 testing, we found no difference in outcomes related to delivery or the neonate among infected women compared to non-infected, nor did severity of infection affect the results. The results indicate that the outcomes of SARS-CoV-2 infection in pregnancy might not be as severe as proposed by previous studies in which more severe cases were included. Testing strategy and how cases are included naturally influence the results and should be considered.

We found that the incidence of termination of pregnancy among SARS-CoV-2 infected women was increased compared to that in non-infected pregnant women, which might be related to universal screening of women admitted to hospital. Further studies are needed to explore a possible relation between late termination of pregnancy and SARS-CoV-2 infection.

## Data Availability

Data in the registry study cannot be shared by the authors but is available upon request from the Danish National Health Data Authority through their research service (https://sundhedsdatastyrelsen.dk/da/forskerservice).
Data from the Danish COvid-19 in pregnancy Database (DCOD) database can be requested by contact to the first author.

## Acknowledgement

The authors would like to thank EasyTrial, who provided their clinical trial management software, used for data management in the DCOD study, free of charge for this study as the company offered free access to the software to COVID-19 research projects.

The authors also wish to thank Professor Marian Knight, DPhil, National Perinatal Epidemiology Unit, Nuffield Department of Population Health, University of Oxford, UK, for her initiation of the International Network of Obstetric Survey Systems study, in which the DCOD participates, and her contribution to the international case report form, on which the DCOD is based.

## Author Contributions

AA, LK, MB, LS, KHR, JJ, LP conceived the registry study, and AA and LK conceived the DCOD study. AA, LK, MB designed and planned the combined study. AA, MB, TP, FP acquired and analysed the data for the cohort study. KW, MI, FJ, ER, CA, IS, TC, JM, LB, BL, AT, MK, BH, LJ, SS, LS, KK, MP, ÅK, MV, DT, MT, LA, AB, AG, CBA, RF, LH, LHv, AS, SR, AA, and LK acquired and managed the DCOD dataset and AA analysed the DCOD data. AA wrote the first draft of the manuscript, MB, LK, TP, LS, KHR and JJ revised the manuscript critically, and all authors approved the final version of the manuscript. All authors accept responsibility for the paper.

### Abbreviations

BMI: Body mass index
CI: Confidence interval
COVID-19: Coronavirus disease
DCOD: the Danish COVID-19 in pregnancy database
DNPR: The Danish National Patient Register
EDD: Estimated date of delivery
GA: Gestational age in completed weeks
HR: Hazard ratio
ICU: Intensive care unit
IQR: Interquartile range
LMP: Date of the first day of the last menstrual period
MiBa: Danish Microbiology Database
NICU: Neonatal intensive care unit
OR: Crude odds ratio
PCR: Polymerase chain reaction
SARS-CoV-2: Severe acute respiratory syndrome coronavirus 2
W: Weeks

## Legends to figures and tables

**Table S1**. Procedure and diagnostic codes used to identify the overall study population in the Danish National Patient Register (DNPR).

**Table S2**. Procedure and diagnostic codes used to identify basic characteristics and outcomes in the Danish National Patient Register (DNPR).

**Table S3**. Definition of pre-existing medical problems and previous pregnancy-related complications in DCOD.

**Table S4**: Additional basic characteristics of SARS-CoV-2 infected women overall and according to severity of infection and all pregnant women in Denmark between March 1 and October 31, 2020.

**Figure S1**: Absolute number of SARS-CoV-2 positive cases and overall incidence per 1000 pregnancies per month in Denmark between March 1 and October 31, 2020.

