## Supplemental tables and figures for "SARS-CoV-2 infection in pregnancy in Denmark – characteristics and outcomes after confirmed infection in pregnancy: a nationwide, prospective, population-based cohort study"

### Figure S1

Absolute number of SARS-CoV-2 positive cases and overall incidence per 1000 pregnancies per month in Denmark between March 1 and October 31, 2020.

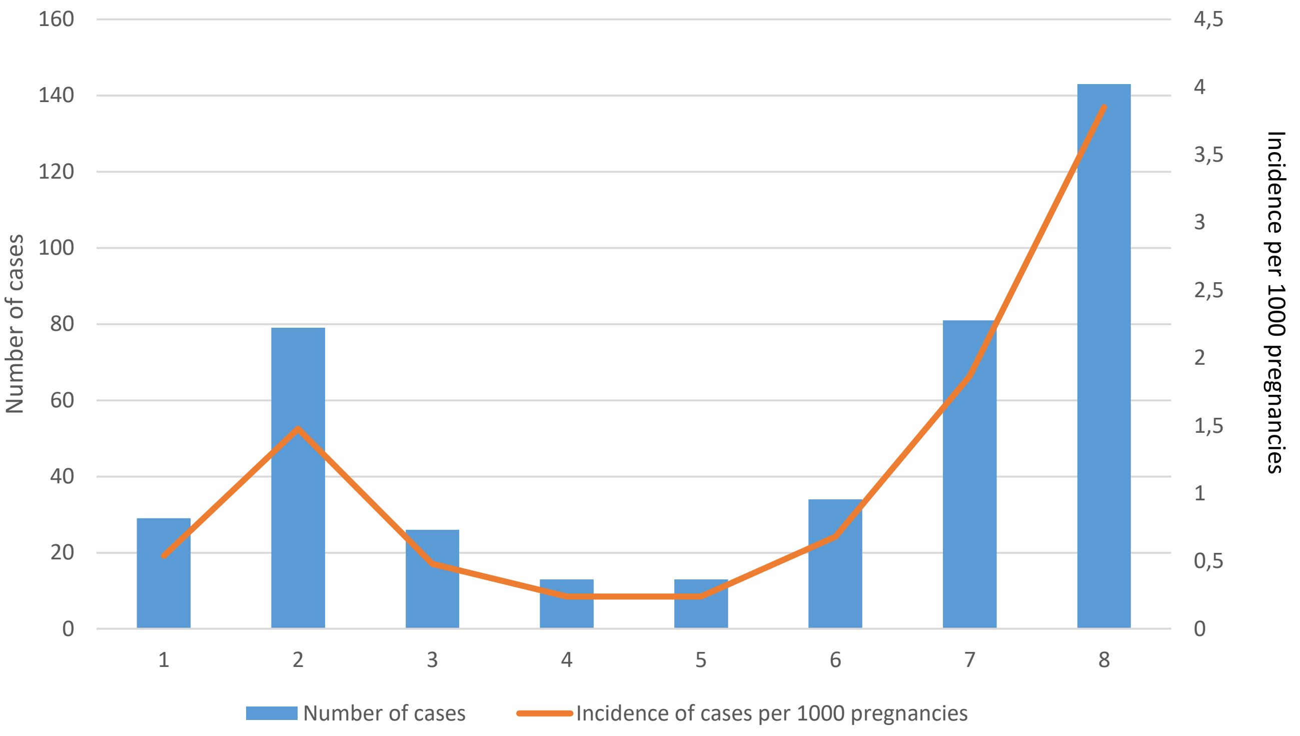

#### Supporting Information Table S1

Procedural and diagnostic codes used to identify the overall study population in the Danish National Patient Register (DNPR).

First, pregnancies were identified in the DNPR if they had a contact with a primary (A) or secondary (B) diagnosis of any of the following codes from March 1 to October 31, 2020 (N = 84 862).

| **Code** | **Description** |
|---|---|
| DZ32* | Confirmed pregnancy diagnoses |
| DZ34*– DZ35* | Pregnancy classification codes |
| DZ36* | Prenatal screening codes |
| DO0*–DO07* | Abortion diagnoses |
| DO200*–DO209* | Bleeding in pregnancy before gestational week 22 diagnoses |
| DO80*–DO84* | Delivery diagnoses |
| UXUD86* | Fetal and placental ultrasound/Doppler procedures |

* indicates that all subdiagnoses/procedures within the overarching code are also included.

Secondly, only pregnancies that could be classified according to the following hierarchy were included in the final register-based pregnancy population. Otherwise, they were excluded.

Hierarchy of classification:

| Hierarchy | Classification | Code | Conditions |
|---|---|---|---|
| 1 | Live birth | DZ38*, DO80*–DO84* | Active personal identification number of the child in the Civil Registration System[a] |
| 2 | Stillbirths/IUFD | DP95*, DO365* | Gestational week ≥22–45 |
| 3 | Miscarriage | DO00*-DO03*, DO08*, DO365* | Gestational week <22 |
| 4 | Termination | DO04-DO07*, BKHD*-BKHD5*, KLCH* | Gestational week <23 |
| 5 | Pregnant at end of follow-up | UXUD86A- UXUD86B, UXUD86N | Gestational age <45 weeks at end of follow-up |

[a]Except births with a record of miscarriage before gestational age (GA) 22 or a termination of pregnancy before GA 23.

Criteria for exclusion from the pregnancy population were:

I. Liveborn child with a missing gestational age (n = 406)

II. Women without a registration of a birth, miscarriage, termination of pregnancy, or ultrasound during follow-up (n = 2965)

III. Women without a registration of a birth, miscarriage, or termination of pregnancy AND a pregnancy of more than 45 gestational weeks at the end of follow-up (n = 187).

#### Supporting Information Table S2

**Procedure and diagnostic codes used to identify basic characteristics and outcomes in the Danish National Patient Register (DNPR).**

| Variable | Code |
|---|---|
| BMI | RDA26*<br>RDA27* |
| Smoking | RDA25* |
| Pre_existing_astma | DO995A<br>DJ45* |
| Hypertension | DO100<br>DI10* |
| Diabetes | DO240<br>DO241<br>DO245 |
| Parity | DZ340<br>DZ348A<br>DZ348B |
| Multiple pregnancy | DO30*<br>(the variable also gets the value 1 if there are twins in the population of children) |
| Eclampsia | DO14<br>DO15 |
| Gestational Diabtes | DO244 |
| Thromboembolic_event | DO882<br>DO223<br>DO225 |
| Pneumonia | DJ12*<br>DJ18* |
| Intensive_care | NABB<br>NABE |
| Death | 90 |
| Termination of pregnancy | DO04*-DO07*<br>BKHD4<br>KLCH03<br>KLCH00 |
| Miscarriages | DO364<br>DO00*-DO03* |
| Ectopic pregnancy | DO00 |
| Total deliveries | D080*-DO84* |
| Vaginal deliveries | DO80*<br>DO81*<br>DO83*<br>DO84* (not DO842)<br><br>If codes for emergency_cd OR |

| | |
|---|---|
| | elective_cd are recorded the variable gets the value 0<br><br>Restricted to deliveries |
| Operative vaginal delivery | KMAE00<br>KMAE03<br>KMAF00<br><br>Restricted to deliveries |
| Emergency Cesarean section | KMCA10A<br>KMCA10D<br>KMCA10E<br><br>Restricted to deliveries |
| Elective Cesarean section | KMCA10B<br><br>Restricted to deliveries |
| Induction | KMAC00*<br>KMAC96A*<br>BKHD20*<br>BKHD20A*<br>BKHD21*<br><br>Restricted to deliveries |
| Total number of children | Derived variable of live birth + IUFD/stillbirths |
| Live birth | pnr!=. and DZ38 |
| IUFD / stillborn | DO364 AND<br>DP95*<br>Restricted to children |
| Neonatal_death | D_status<br>D_doddato<br>D_foddato<br>Restricted to live born children |
| Apgar score 5 min. | DVA00-DVA10<br>DVAXX<br>Restricted to live born children |
| Arterial pH | RDA46*<br>Restricted to live born children |
| Venous pH | RDA47*<br>Restricted to live born children |

* indicates that all subdiagnoses/procedures within the overarching code are also included.

#### Supporting Information Table S3

**Definition of pre-existing medical problems and previous pregnancy-related complications in DCOD.**

*Pre-existing medical problems* comprised cardiac, renal, endocrine, psychiatric, hematologic, and autoimmune diseases; cancer; and HIV.

*Previous pregnancy-related complications* included thromboembolic events, eclampsia, habitual abortion, preterm birth, mid-trimester loss, neonatal death, stillbirth, baby with major congenital abnormality, small/large for gestational age infant, infant requiring intensive care, puerperal psychosis, placenta praevia, gestational diabetes, placental abruption, postpartum haemorrhage requring transfusion, surgical procedure during pregnancy, hyperemesis and dehydration requiring admission, ovarian hyperstimulation syndrome or severe infection.

**Table S4**: **Additional basic characteristics of SARS-CoV-2 infected women overall and according to severity of infection and all pregnant women in Denmark between March 1 and October 31, 2020.**

| | | SARS-CoV-2 infected pregnant women identified in DCOD | SARS-CoV-2 infected pregnant women / pregnancies identified in the national registers | Non-infected pregnancies identified in the national registers | SARS-CoV-2 infected vs. non-infected pregnancies | | SARS-CoV-2 infected women identified in DCOD | | |
|---|---|---|---|---|---|---|---|---|---|
| | | | | | DCOD SARS-CoV-2 cases | Register SARS-CoV-2 cases | Women requiring admission to hospital due to COVID-19 symptoms | Women with uncomplicated SARS-CoV-2 infection* | Women requiring hospital admission due to COVID-19 symptoms vs. women with uncomplicated SARS-CoV-2 infection* |
| | | N = 418 | N = 420 | N = 82 262 | $OR_D$ (95% CI) | $OR_R$ (95% CI) | N = 23 | N = 395 | $OR_{DCOD}$ (95% CI) |
| **Basic characteristics** | | | | | | | | | |
| Hypertension | | < 3 | <5 | 719 (0.9) | NA | NA | NA | NA | |
| Diabetes | | 3 (0.7) | <5 | 492 (0.6) | 1.20 (0.39–3.75) | NA | NA | NA | |
| Women in employment | | 314 (79.3) | NA | NA | | | 16 (72.7) | 298 (79.7) | 0.68 (0.26–1.80) |
| | Unknown | 22 (5.3) | | | | | 1 (4.3) | 21 (5.3) | |
| **COVID-19 characteristics** | | | | | | | | | |
| Test type | PCR or antigentest | 399 (97.3) | NA | NR | | | 23 (100) | 376 (97.2) | 1.03 (1.01–1.05) |
| | IgG in serum | 11 (2.7) | NA | NR | | | 0 (0) | 11 (2.8) | NR |
| | Unknown | 8 (1.9) | | | | | 0 (0) | 8 (2.0) | NR |
| Administration of anti-virals | | < 3 | NA | NR | | | < 3 | 0 (0) | NA |
| | Unknown | 7 (1.7) | | | | | 1 (4.3) | 6 (1.5) | |
| **Pregnancy characteristics** | | | | | | | | | |
| Previous pregnancies | Multigravida | 275 (66.9) | NA | NA | | | 17 (73.9) | 258 (66.5) | 1.43 (0.55–3.71) |
| | Unknown | 7 (1.7) | | | | | | 7 (1.8) | |
| Previous pregnancy related complications among multigravid women[†] | | 60 (21.9) | NA | NA | | | 3 (17.6) | 57 (22.2) | 0.75 (0.21–2.71) |
| | Unknown | 1 (0.4) | | | | | 0 (0) | 1 (0.4) | |

Data are presented as count (%) or median (interquartile range)

$OR_D$ was estimated using 2 x 2 contingency tables. $OR_R$ was estimated with univariate logistic regression.

$OR_{DCOD}$ was estimated using logistic regression.

* Uncomplicated COVID-19 included cases where women did not require admission to hospital due to COVID-19 symptoms. Includes women in home isolation or women admitted for obstetric reason within 14 days of first positive COVID-19 test or symptoms.

† Previous pregnancy-related complications include thromboembolic event, eclampsia, habitual abortion , preterm birth and mid trimester loss, neonatal death, stillbirth, baby with major congenital abnormality, SGA, LGA, infant requiring intensive care, puerperal psychosis, placenta praevia, GDM, placental abruption, PPH requiring transfusion, surgical procedure during pregnancy, hyperemesis or dehydration requiring admission, OHSS or severe infection.

BMI Body mass index; DCOD Danish COVID-19 in pregnancy database; GA Gestational age; NR Not relevant; NA Not available
